# Psychosocial risk factors for injury in performing artists: A scoping review of screening and predictive instruments

**DOI:** 10.1101/2025.04.04.25325124

**Authors:** Róisín Cahalan, Caoimhe Barry Walsh, Orfhlaith Ni Bhriain, Breandán de Gallaí, Hannah Fahey de Brún, Michele Pye, Rose Schmieg

**Affiliations:** School of Allied Health, University of Limerick, Limerick, Ireland; Physical Activity for Health Research Centre, University of Limerick, Limerick, Ireland; Irish World Academy of Music and Dance, University of Limerick, Limerick, Ireland; Shenandoah University, Division of Athletic Training, Shenandoah University, Virginia, United States; Department of Physical Therapy, College of Nursing and Allied Health Science, Howard University, Washington DC, United States

## Abstract

The performing arts is a diverse collection of disciplines. However common to all disciplines is an elevated risk of injury and an array of biopsychosocial risk factors. While screening for physical risk factors is common practice, and largely routine, psychosocial screening for injury in performance artists (PAs) is less well established. This scoping review aimed to systematically map, and report the suitability of, instruments used to screen or assess psychosocial risk factors for injury in non-recreational adult performing artists (PA)s. This scoping review was conducted in accordance with the Joanna Briggs Institute Evidence Synthesis guidelines. Twelve databases relating to performance, health, medicine, kinesiology, and sport were searched. Studies that investigated associations between psychosocial factors and injury in non-recreational (professional, pre-professional, full-time collegiate students, elite competitive) adult PAs were eligible. Fifty-one studies of 7,457 participants (25 of musicians (n=4,505 (60.5%)); 24 of dancers (n=2,680 (35.9%)); 1 of vocalists (n=225 (3.0%)), and 1 of circus performers (n=47 (0.6%))) met the inclusion criteria. Most participants were professional PAs (n=4,547 (61.0%)), followed by collegiate PAs (n=1,424 (19.1%)), and mixed professional, pre-professional, elite competitive, and collegiate PAs (n=1,486 (19.9%)). A total of 90 distinct instruments evaluating 45 different psychosocial factors were identified. Stress, anxiety, depression, perfectionism and coping were the factors most frequently investigated. Stress was commonly reported across all PA cohorts. Just 19 (21%) instruments were psychometrically appropriate for the cohort being measured. Many other instruments were valid/reliable in patient, sporting or general population cohorts, but not in PAs. There is a common link between psychosocial risk factors and injury in non-recreational adult PAs. Screening programmes should incorporate comprehensive evaluations of these factors. Instruments appropriate for the cohort being investigated should be used. The development and/or validation of instruments for use across all PAs should be considered.

## Introduction

The performing arts is a broad church that encompasses diverse disciplines where artists use their voices, bodies, or inanimate objects to convey artistic expression(1). This includes, but is not limited to, music, dance, song, circus and theatre arts. Although predominantly aesthetic activities, many performing artists (PAs) exist at the nexus between the artist and athlete. Significant physical and psychological ability and reserves are required to withstand the rigours of the highly repetitive nature of performance training and the threat of pain and injury(2).

The occurrence of injury across various disciplines in the performing arts is well documented. A recent review of dancers from various genres and at different levels (recreational, student elite and professional dancers) reported that injury in dance remains a concerning issue with a paucity of high-level evidence and inconsistency in research methods (3). Injury incidence in dancers has been found to range from 0.16 to 4.6 per 1000 hours of exposure (4, 5), with point and period prevalence of 54.8% (6) and 280% (7) respectively, reported in the literature. Similarly in musicians, high levels of pain, weakness and other musculoskeletal disorders have been recorded (8, 9). A recent systematic review of adult musicians reported a lifetime injury prevalence of 46 – 90% in this cohort (10). In vocalists, phonotrauma is largely understood to stem from cumulative vocal fold tissue damage and/or response to persistent tissue inflammation (11). An overall career prevalence of self-reported dysphonia (hoarse, raspy, strained voice) in singers of 46.09% has been reported in the literature (12). Furthermore, it is estimated that up to 29% of attendees at voice clinics in the United States are professional vocalists, even though they represent less than 1% of the national work force (13).

Interestingly, there is considerable overlap in many of the identified causes of injury across the performing arts. Much of the research in this area focuses on physical causes of injury, with factors including overtraining, excessive load and under-recovery identified in studies across music, song and dance respectively (14-16). A study exploring injury in dancers and musicians identified a host of biomechanical issues, poor ergonomics and suboptimal technique as key factors driving injury in both groups (17). Prior injury is also noted as an important risk factor for future injury across many areas of the performing arts (18-20).

There is also a growing appreciation of the association between psychosocial factors and injury across various fields in the performing arts (21-23). Music performance anxiety and high levels of stress were found to be associated with an increased level of musculoskeletal disorders in a systematic review of professional and pre-professional instrumentalists (19). Emotional exhaustion, poor self-efficacy and fatigue were related to an increased injury risk in a cohort of circus artists,(24) while in vocalists, an association between personality traits/facets related to happiness, dominance and caution, and phonotrauma were reported (15). A multitude of factors including stress, psychological distress, disordered eating, poor coping, suboptimal sleep, personality, and social support have also been found to be associated with increased injury risk or adverse injury outcomes in dancers (25).

In addition to shared injury experiences, PAs have much in common regarding the identity of the artist, and attitudes to performance when ill or injured. Continuing to perform when in pain or injured is a common finding in dancers (26), musicians (27) and vocalists (28). These authors cite reasons that range from the more benign, such as a mild injury that is likely to resolve itself, to more sinister motivations including stigma associated with being injured, a culture of concealment, and fear of losing roles, work and status. Additionally, the experience of being seriously injured can be devastating for the PA, whose identity is intimately entwined with their art (29, 30). The extent to which the PA experiences these challenges to their identity, and repercussions therein are known to be impacted by many factors including personality traits, coping strategies, and quality of social support (31).

While the increasing focus on psychosocial drivers of injury is encouraging, injury screening practises are still overwhelmingly concerned with assessing physical risk factors such as strength, range of movement, fitness and other biomechanical markers (2). Recent efforts to develop a holistic screening instrument for PAs including physical and psychological evaluations specific to individual cohorts are promising (32), but large-scale studies are lacking. Thus, instruments and techniques to evaluate physical risk factors in PAs are well established and psychometrically sound in many cases. It is unclear whether many of the instruments used to evaluate psychosocial factors in PAs are equally robust.

It is our contention that, despite the heterogeneous physical demands of PAs, the psychosocial profile of injured PAs is similar across disciplines. There may, therefore, be an opportunity for common assessment instruments to screen for and identify psychosocial injury risk across disciplines. An important initial step is to catalogue both the instruments used for this purpose, the specific PA cohorts they have been used with, and the psychosocial risk factor that they evaluate. This scoping review, therefore, aimed to systematically map and report the suitability of instruments used to screen or assess psychosocial risks for injury in non-recreational PAs.

## Methods

### Protocol and registration

Owing to the heterogeneity of the available literature, a scoping review was identified as the most appropriate methodology to address the aims of this study (33). This scoping review was conducted in accordance with the Joanna Briggs Institute (JBI) Evidence Synthesis guidelines (34) and Preferred Reporting Items for Systematic Reviews and Meta-Analyses extension for Scoping Reviews (PRISMA-ScR) (35). A scoping review protocol in accordance with the Joanna Briggs Institute was registered on the Open Science Framework (https://osf.io/sjntx/).

### Inclusion criteria

- Original peer-reviewed research, clinical practice guidelines reporting assessment or evaluation instruments (including surveys, questionnaires) of psychosocial factors associated with injury in PAs. For conciseness, the term “injury” will be used in this text, but references to pain, physical disorders, wounds and ailments in the literature were also included in search parameters. (Tab 1). Pertinent systematic reviews in the area were screened for studies meeting the inclusion criteria.
- Studies involving adult (18 years and over) dancers, musicians, singers, circus artists, or other physical performance (e.g. theatre actors).
- Non-recreational PAs, including professional, pre-professional, full-time University (or equivalent) performing arts students, or otherwise described elite performers (such as competitive at highest possible level in that genre) were included. Studies of mixed (recreational and non-recreational) artists were only included if data pertaining to non-recreational performing artists could be extracted.
- English language, human research studies published from the date of inception of the database.

**Table 1:** Key concepts informing search strategy.

| **Concept A** | **Concept B** | **Concept C** | **Concept D** | **Concept E** |
| --- | --- | --- | --- | --- |
| **Risk** | **Activity** | **Population** | **Injury** | **Measurement** |
| Psycho-social | Performing artists | Adult | Musculoskeletal injury | Instrument or tool |
| Stress | Dance | Professional | Pain | Survey |
| Anxiety | Vocalist/singer | Pre-professional | Wound | Questionnaire |
| Catastrophis(z)ing | Musician/instrumentalist | Full-time Student | Disorder | Assessment |
| Depression | Theatre actor | Elite | Ailment | Screening |
| Cognitive/mental | Circus |  |  |  |

### Exclusion criteria

- Studies including instruments which assessed/measured solely physical factors including load and physical fatigue.
- Instrument was not used to assess a relationship/association with injury.
- Qualitative (non-instrument based) reports of psychosocial issues/risks.
- Studies of recreational PAs.
- Study protocols as associations between variables are not recorded, case reports, abstracts, conference proceedings and other secondary research studies.
- Studies of PAs under the age of 18 years.

### Information sources

Database searches were conducted in November 2024 and limited to the following databases: Web of Science; EMBASE; Cochrane Database of Systematic Reviews; EBSCO (CINAHL Ultimate, MEDLINE, SPORTDiscus, PsycINFO); PubMed; Elsevier (ScienceDirect; Scopus); ProQuest Performing Arts Periodical Database, Dissertations); SAGE; JSTOR; and PEDro: the Physiotherapy Evidence Database. A concept framework approach was adopted to design the search strategy around the eligibility criteria for each database (Table 1).

### Study Selection and Screening

In conjunction with the faculty librarian at the host institution of the lead author (RC), a pilot search strategy was developed and tested. Subsequently, appropriate subject headings and keywords were identified for the final search strategy and adapted for each database. All pertinent records identified in the search were collated into Endnote X9.3.3 (Clarivate Analytics, PA, USA) and citation details were imported into the Covidence reference management system (Covidence; Covidence Melbourne, Australia). Duplicates were removed and titles and abstracts were screened by two independent reviewers (RC & CBW) for assessment against the inclusion and exclusion criteria previously outlined. Potentially relevant papers were retrieved and the full text assessed in detail against the inclusion and exclusion criteria by pairs of independent reviewers from the authorship team (ONB, MP, HDB & BDG). Just one disagreement arose between the reviewers at this stage of the selection process which was adjudicated by an additional authorship team member (RS). Full texts of selected papers were hand searched for additional relevant papers referred to in the appendices. This resulted in the addition of a further nine studies.

### Data charting, collection and extraction

Data charting, collection, and extraction for this scoping review followed a systematic and transparent process to ensure comprehensive capture of relevant information. A standardized data extraction form based on the JBI Manual (34) (Appendix 10.1) was used by all members of the research team who completed data extraction. The form was amended to detail key information from eligible studies including included study characteristics (author, year, country of origin), characteristics of instruments (name, purpose, risk factor measured), study design, study population (profession, sex, age), settings, the psychometric performance of instruments where available, and any findings related to relationship between psychosocial factor and injury. Data extraction was completed by authors RC and CWB and reviewed for accuracy by the remaining members of the authorship team. Disagreements were adjudicated by RS.

## Results

Initial searching identified 2726 articles for screening. Following title and abstract review, 144 full texts were assessed against the inclusion and exclusion criteria. The initial title and abstract (n=270, 10%) and follow-up full text (n=29, 20%) screening pilots of these articles had an agreement of >85% between reviewers. A total of 51 studies met the inclusion criteria following full-text review and were included in the scoping review (Fig 1).

**Fig 1:**
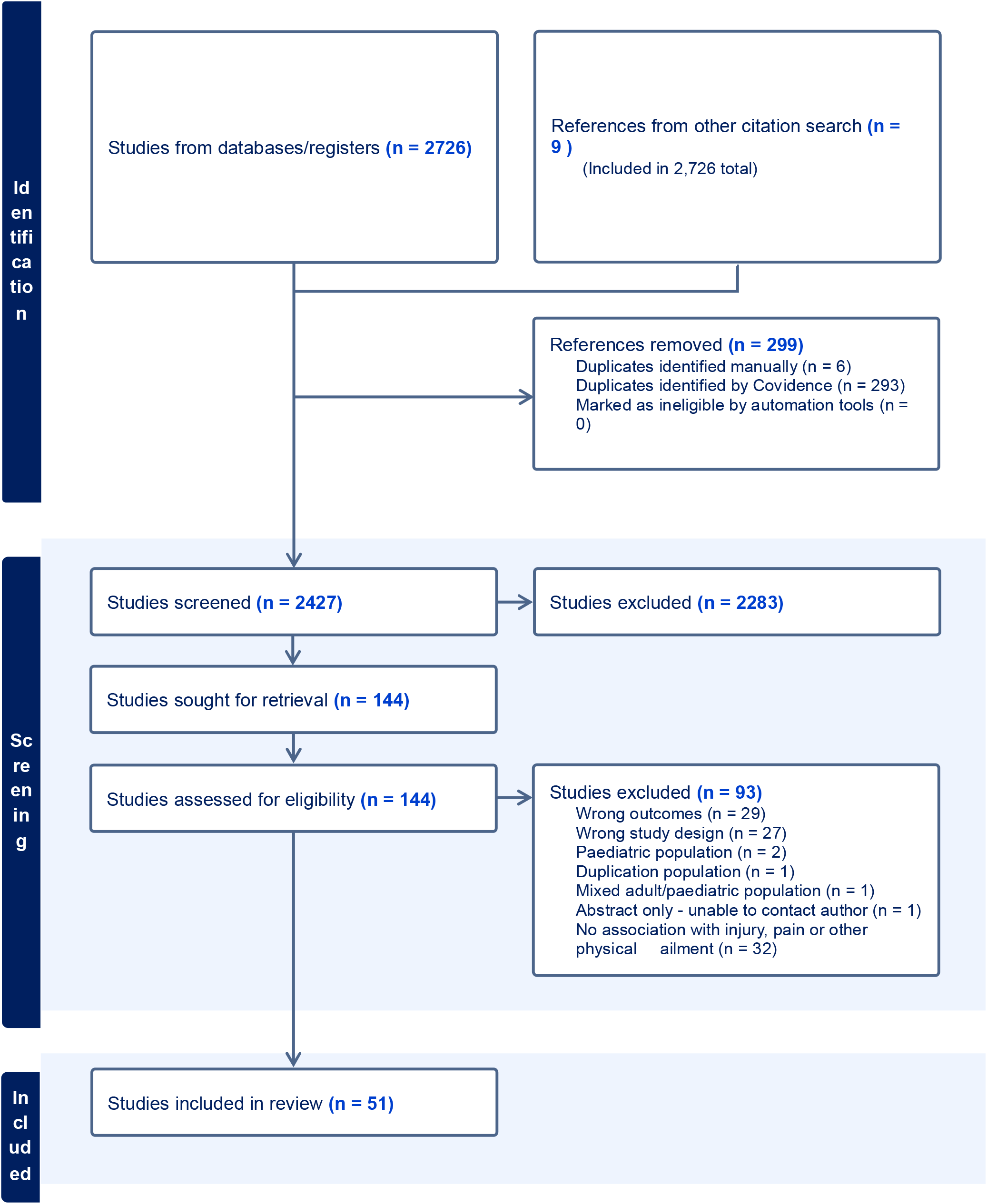
PRISMA Flow diagram of studies screened and included in this review.

The 51 articles in this review included 25 studies conducted in musicians, 24 in dancers, and a single study each investigating vocalists and circus performers. No studies of theatre performers or other PAs met the criteria for inclusion in this review. The studies were conducted primarily in Europe (n=29 (56.9%)), and North America (n=13 (25.5%)), with four (7.8%), three (5.9%) and two (3.9%) studies each from Australia, South America and Asia respectively. Fig 2 outlines the geographical location and cohort focus of the included studies.

**Fig 2:**
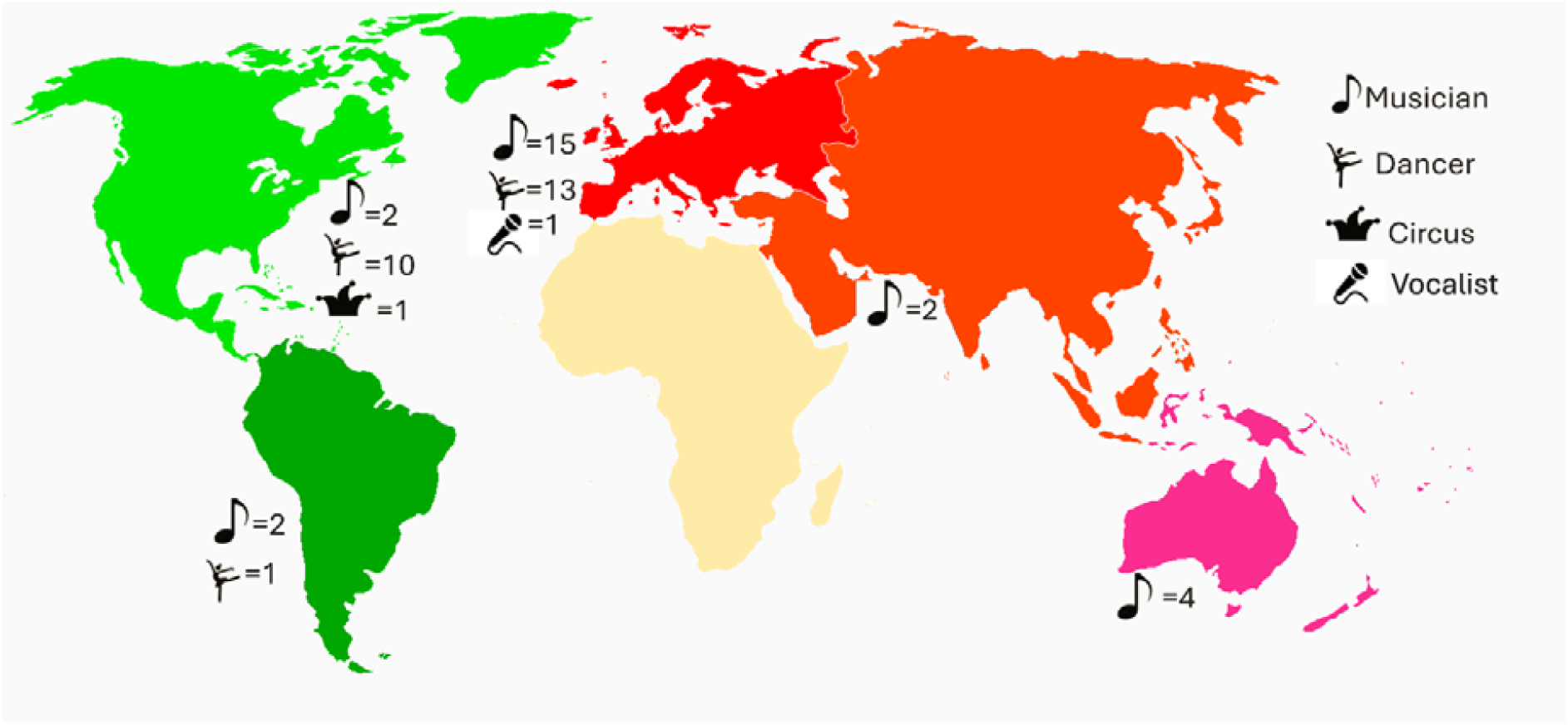
Focus and geographic distribution of studies by continent.

A total of 7,457 participants were included in this review (male: n=3,215 (43.1%); female: n=4,242 (56.9%); unrecorded: n=11 (0.1%). Participants were drawn from musician (n=4,505 (60.5%)), dancer (n=2,680 (35.9%)), vocalist (n=225 (3.0%)) and circus (n=47 (0.6%)) populations respectively. Most participants were professional PAs (n=4,547 (61.0%)), followed by collegiate PAs in full-time programmes of study in their respective arts (n=1,424 (19.1%)). The balance of participants was drawn from six studies of mixed professional, pre-professional, elite competitive, and collegiate PAs (n=1,486 (19.9%)). Study characteristics are outlined in S1 Table.

A total of 81 distinct instruments were used by authors to evaluate various psychosocial domains (S2 Table). Additionally, nine bespoke instruments including visual analogue scales and Likert scales were created and used by authors to interrogate various psychosocial criteria of interest. A number of established instruments were used in multiple studies with the most popular including the Kenny Music Performance Anxiety Inventory (K-MPAI)(36), and various subscales of the Short-Form Health Survey (SF36)(37), which were both used in five studies. The State-Trait Anxiety Inventory (STAI)(38) was used in four studies and the Hospital Anxiety and Depression Scale (HADS)(39) and Profile of Moods State (POMS)(40) questionnaire were both used in three studies.

Of the 81 instruments used in these studies, the psychometric properties of just 19 were reported in the cohort under investigation. This included 13 instruments used in dance cohorts and six in musicians, one of which has also evaluated in a cohort of vocalists (S2 Table). Typically, acceptable levels of internal consistency (test-retest reliability) were established for the use of these instruments in the pertinent cohort (S2 Table). A further two instruments; the Dancer Injury Profile Questionnaire (41) and the University of North Texas Musician Health Survey (42), which were designed specifically for dancers and musicians respectively were used, however psychometric analysis of these instruments was unavailable in the literature. Finally, an abbreviated version of the First Multidimensional Perfectionism Questionnaire, (43) an instrument identified in this review, was also found to be reliable in a cohort of elite opera singers (44). The reliability and/or validity of most instruments used in the included studies may have been established in other cohorts including athletes, patient groups or the general public, but not specifically in the cohort of PAs under consideration.

A total of 45 different psychosocial factors were explored (Tab 3), with many studies assessing multiple factors. There were also many outcome measures which assessed more than one psychosocial factor concurrently (e.g. Depression Anxiety Stress Scale). This rendered it impractical to report percentages of each psychosocial factor explored. The most investigated psychosocial factors in the reviewed studies were anxiety (generalised, performance, competition, social and workplace), depression, coping and perfectionism. These factors were consistently reported in studies of dancers and musicians, with stress presenting as an issue in the single studies involving vocalists and circus performers. Of all studies included in this review, just four studies – all in dancers - reported no relationship between the psychosocial factor investigated, and injury in participants (45-48). Details of associations/correlations reported between psychosocial factor and injury are detailed in S2.

**Tab 3:** Psychosocial factors investigated in reviewed studies.

**Tab 3: Psychosocial factors investigated in reviewed studies.**
|  |  |  |  |  |
| --- | --- | --- | --- | --- |
| Adaptability | Coping | Holistic wellness | Multi-dimensional strain | Self-esteem |
| Alcohol addiction | Core self-evaluation | Hypervigilance | Mental fatigue | Self-regulation |
| Anxiety | Depersonalisation | Inter-personal sensitivity | Pain-related performance | Sleep disturbance |
| Body image | Depression | Job satisfaction | Passion | Sleep quality |
| Burnout | Disordered eating | Job stress | Passion for dance | Social phobia/anxiety. |
| Catastrophising | Dissociation | Legacy of trauma | Perfectionism | Social support |
| Childhood adversity | Emotional regulation | Life stressors | Performance anxiety | Stress |
| Competence and control | Fear-avoidance behaviours | Mental disorder | Personality traits | Temperament |
| Competition anxiety | Fear-avoidance beliefs | Mood | Resilience | Tension |

**Note:** Percentages of each factor evaluated is not reported due to a range of instruments that reported multiple factors concurrently.

## Discussion

This scoping review has collated a comprehensive list of the instruments used to evaluate relationships between a variety of psychosocial factors and the risk of injury, pain or other physical problems in non-recreational adult PAs. The extensive number of studies that met the inclusion criteria for this review underscore the central and consequential role that psychosocial factors such as stress, anxiety, depression and others play in the physical performance and overall wellbeing of dancers and musicians in particular, whilst further studies are required to establish the extent of this relationship in vocalists and circus performers. This scoping review also furthers the hypothesis that PAs from diverse genres have much in common regarding the psychosocial factors that influence, and potentially mitigate injury risk.

While this review has comprehensively illustrated the relationship between psychosocial factors and injury in dancers and musicians, there exists a broad swathe of literature highlighting the existence of these factors in vocalists. The presence of performance anxiety (49), neuroticism (50) occupational stress and perfectionism (44) have all been identified in the literature. Many of these studies did not explore a relationship with injury and thus were not included in this review. Similarly in professional circus performers, levels of depression, anxiety, stress and overall mental health have been found to be worse than normative scores in the general population, but mediated by comparatively higher resilience (51). Comparable findings relating to higher than normal general population levels of moderate psychological stress have also been reported in a cohort of 92 circus student-artists (52). Qualitatively, the link between adverse psychosocial factors and injury has also been reported in numerous studies of PAs across the various professions. In a cohort of professional orchestral musicians, participants identified a direct relationship between injury and factors including performance, workplace and relationship stress (53). Qualitative research in elite dance has identified how the drive for perfection and the pressures of competition and oppressive power dynamics may push dancers into pain and injury, which are then normalised due to a subculture of perseverance and concealment (54). Similarly, vocalists have self-reported perceived links between performance anxiety, family pressures, depression and a range of somatic problems (55).

It therefore follows that it is critically important to design appropriate screening protocols to identify and mitigate the presence of key psychosocial drivers of injury in these PAs. As mentioned, a disproportionate focus on screening of physical traits persists despite the ample evidence of the importance of psychosocial factors. For instance, protocols that solely consider phonatory agility, strength, and stamina in vocalists (56) or focus on range of movement (ROM), hypermobility or balance in dancers (57) In some cases, efforts have been made to consider a small number of psychosocial factors, such as an evaluation of coping in a ballet dance cohort, but once again the vast majority of the protocol referred to physical elements such as flexibility or balance (58). Given the array of psychosocial issues identified in this review, and their association with injury risk, a more comprehensive approach is required.

The novel “Dancers, Instrumentalists, Vocalists, and Actors” (DIVA)(32) screening protocol considers a more diverse array of injury risk factors including a checklist of psychosocial factors, and the KMPA-I questionnaire as well as general health, activity details, orthopaedic and vocal/audiology items. This is a welcome development, as it may be used and adapted by PAs from a variety of backgrounds. It may, however, inadequately address important factors such as perfectionism, burnout, coping and other factors which were identified in this review. Work by Rousseau and colleagues in the development of a multi-dimensional, risk-based injury screening model for musicians is encouraging (59). This evidence-based, stakeholder-informed project considered the impact of approximately 15 different psychosocial factors in addition to potential sources of risk including individual characteristics, biomechanics, posture, life habits, workload and physical condition. In doing so, a truly comprehensive evaluation of the holistic risk and vulnerability profile of the musician is possible. It is equally important however to ensure that appropriate instruments are used to evaluate the presence of the psychosocial factor in question.

In this review, the vast majority of instruments used by authors, had not been psychometrically tested in the cohort of interest. Assumptions made regarding the transferability of instruments designed for use in patients, athletes or the general public have not been confirmed in all cases. However, there are plentiful instruments identified in this review that have been designed specifically for various types of PAs, such as the KMPA-I which is appropriate for use in musicians (36) and vocalists (44), and the Passion for Dance Scale in dancers (41). Additionally, there are a host of other instruments not discovered in our search that are appropriate for specific performing arts cohorts including the Scale of Coping with Pain for Dancers (COPAIN-Dancer)(60) and the Mazzarolo Music Performance Anxiety Scale (M-MPAS)(61). These instruments have been developed relatively recently and may not have been available to the authors of many studies included in this review. It may also indicate an encouragement move towards the development of more PA-specific tools. Similar bespoke instruments for vocalists and circus performers are lacking. However, given the shared experiences of PAs from disparate genres, research should explore if these instruments are suitable, or can be adapted, for use in these under-served groups.

## Limitations

Although beyond the scope this review, many of the studies were cross-sectional, retrospective studies and therefore of lower methodological quality. A number of studies used instruments that evaluated multiple psychosocial factors concurrently, making it impossible to accurately report specific percentages of factors evaluated in the studies in this review. The exclusion of non-English language studies may have led to the omission of information relevant to this review.

## Conclusion

Psychosocial issues are commonplace and associated with pain, injury and other physical problems in non-recreational PAs. A plethora of instruments have been used to evaluate this relationship, many of which have not been established as valid and/or reliable in the population of interest. In addition to the evaluation of physical risk factors for injury, screening protocols should commit to comprehensive evaluation of a diverse array of potential psychosocial factors and measure these using appropriate instruments. Importantly, there is a shared experience of psychosocial problems in PAs which transcend profession. Future research should focus on the development and/or validation of instruments that may be used across the entire PA community. Particular focus should be on instruments to investigate the presence of anxiety, stress, depression, perfectionism and coping in PAs.

## Supporting information

Supl 1: Included study details

Supl 2: List of instruments

Supl 3: Search strings

PRISMA Check list

## Data Availability

All relevant data are within the manuscript and its Supporting Information files.

