## Supplementary material for "Psychosocial risk factors for injury in performing artists: A scoping review of screening and predictive instruments": Supl 1: Included study details

| Study details | Nature of study | Participant details | Setting | Biopsychosocial instruments | Findings regarding biopsychosocial factors and injury. |
|---|---|---|---|---|---|
| Ackerman et al.; 2014, Australia(1) | Cross-sectional | Professional orchestral musicians.<br>N = 377 M: 184; F: 192; Unknown: 1. Mean (SD) Age: 42.1 (10.2) | Eight major Australian orchestras | Kenny Music Performance Anxiety Inventory (K-MPAI)<br>State-Trait Anxiety Inventory (STAI-T).<br>Social Phobia Inventory (SPIN)<br>PRIME-MD Patient Health Questionnaire (PRIME-MD PHQ)<br>Anxiety and Depression Detector (ADD)<br>Core Self Evaluation Scale (CSE)<br>Alcohol Use Disorders Identification Kit (AUDIT) | Interactive effects between psychological well-being, PRMDs, and pain. Linear relationship between increasing TPP and MPA scores for females. Higher PRMD severity associated with higher K-MPAI scores. Pain frequency, pain intensity, and pain severity all associated with depression. Risk Factors: MPA, depression, social anxiety, trauma/PTSD, low self-efficacy |
| Ackerman et al.; 2011, Australia(2) | Cross-sectional | Skilled collegiate flute players. N = 20 M: 3; F: 17. Mean (SD) Age: 23.05 (3.76) | Sydney Conservatorium of Music | Bespoke flautists' health questionnaire (location, duration and intensity of PRMDs experienced, practitioners consulted, perceived reasons for injury) | Numerous risk factors attributed to PRMDs including long hours of practice, poor posture, insufficient rest and performance anxiety. |

| | | | | | |
|---|---|---|---|---|---|
| Adam et al.; 2004, Germany(3) | Cross-sectional | Professional ballet dancers. N = 54 M: 24; F: 30. Mean (SD) Age: 26.59 (6.2) | German ballet company and visiting soloists from other European companies | Cohen Perceived Stress Scale<br>Social Support Appraisals Scale<br>Profile of Mood States (POMS)<br>Pittsburgh Sleep Quality Index (PSQI) | Significant positive correlation between perceived stress and time missed due to injury (r = .60, p < .001).<br>Significant positive relationship between injuries and negative mood states: tension-anxiety (r = .41, p < .001), depression-dejection (r = .41, p < .001), anger-hostility (r = .32, p < .01), confusion-bewilderment (r = .44, p < .001), fatigue-inertia (r = -.26, p < .05).<br>Percentage of time injured negatively associated with social support (r = -.29, p < .02).<br>Percentage of time injured positively correlated with daytime sleepiness (r = .50, p < .001) and delayed sleep onset (r = .44, p <.001) |
| Ahlberg et al.; 2019, Finland(4) | Cross-sectional | Symphony orchestra musicians. N=488. M: 255 (52.3%); F: 233 (47.7%). Mean (SD) Age: M: 47.7 (10.3); F: 43.4 (9.8). | Finnish symphony orchestras | Bespoke questionnaire adopting elements of pre-existing instruments assessing sleep, stress, and oro-facial pain (including perceived quality of sleep, possible sleep bruxism, stress experience and oro-facial pain) | Current oro-facial pain significantly associated with disrupted sleep (p = 0.001), frequent sleep bruxism (p < 0.001) and frequent stress (p = 0.002). |
| Amorim & Jorge; 2016, Portugal(5) | Cross-sectional | Professional or semi-professional violinists. N=93; M: 51, F: 42. Mean (SD) Age: 33.4 (12.8) | Orchestras and conservatories around Lisbon, Portugal | Fonseca Anamnestic Questionnaire.<br>Kenny Music Performance Anxiety Inventory (K-MPAI). | Statistically significant association between presence of TMD and high MPA levels (P < 0.001).<br>Most anxious violinists were six times (95% confidence interval 2.51-15.33; p<0.001) more likely to report TMD symptoms. |

| | | | | | |
|---|---|---|---|---|---|
| Austen et al.; 2024, United Kingdom(6) | Cross-sectional pilot study | Music conservatoire students. N = 16. M=4, F= 12. Mean (SD) Age: 23.63 (2.99) | Music conservatoires in UK and Ireland. | Musculoskeletal Pain Intensity and Interference Questionnaire.<br>Generalised Anxiety Disorder (GAD-7).<br>Patient Health Questionnaire 9 (PHQ-9). | Pain Interference: anxiety and depression delta scores were significantly correlated for the exercise group ($p <.05$).<br>Anxiety and pain intensity delta scores were significantly correlated in both groups (exercise and waitlist control) ($p<.05$). |
| Ballenberger et al.; 2018, Germany(7) | Prospective Cohort | Music and non-music university students. N = 63. Musicians: 33 (52.4%). M:22(66%); F: 11(33%) Mean (SD) Age: Musicians: 20.79 (2.43); | University of Applied Sciences Osnabruck, Germany | Stress and Coping Inventory (SCI).<br>Short-Form Health Survey (SF36). | Risk factors for MHC included stress symptoms ($p=0.02$); reduced emotional functioning ($p=0.04$); and decreased positive thinking ($p = 0.06$). |
| Benoit-Piau et al.; 2024, Canada(8) | Prospective Cohort | Professional and pre-professional dancers. N = 88. M: 20 (23%); F: 67 ((76%); Unknown: 1 (1%) Mean (SD) Age: 20.7 (4.2) | Fourteen dance schools and companies. | Passion Scale (PS).<br>Kenny Music Performance Anxiety Inventory (K-MPAI). | A higher level of obsessive passion was associated with a higher incidence of MDEs causing an interruption of dance activities ($\beta = 0.264$, $p = 0.022$). |
| Berlet et al.; 2002, United States(9) | Prospective study | Professional dancers. N = 15. M= 6; F= 9 ;Mean (SD) Age: M: 28.83 (3.31); F 26.89 (2.98). | Orthopaedic Foot and Ankle Centre in Columbus, Ohio | SF36 General Health Status Survey | No significant correlations between injury rate and lifestyle variables (emotional or mental health factors). |
| Byhring & Bo; 2002, Norway(10) | Prospective cohort study | Professional ballet dancers. N=41; M = 14; F=27; Mean age 26.7 (Range 19-41). | Norwegian National Ballet | Bespoke questionnaire including questions related to work stress, job influence, previous injuries, training routines and organisational and environmental factors | No significant association between negative stress, influence at work and musculoskeletal injuries. |

| | | | | | |
|---|---|---|---|---|---|
| Cahalan et al.; 2015, Ireland(11) | Cross-sectional | Professional Irish dancers, elite competitive Irish dancers, and collegiate Irish dancers. N = 104. M = 30; F = 74 Mean (SD) Age: F: 21.2 (3.1); M:23.0 (4.0); | Multiple dance settings across Ireland. | Coping Strategies Questionnaire (CSQ). Profile of Mood States (POMS), Depression Anxiety Stress Scale (DASS). Athletic Coping Skills Inventory (ACSI-28). Sick, Control, One stone, Fat, Food (SCOFF) questionnaire. | Factors significantly associated with being in the "Significantly Injured" group included: higher number of subjective health complaints (p=0.001); higher number of psychological complaints (p=0.036); low mood (p=0.01); heightened catastrophising (p=0.047). No association was found between social support, performance anxiety and injury. |
| Cahalan & O'Sullivan; 2013, Ireland(12) | Retrospective cross-sectional | Professional Irish dancers. N = 178: M:67; F:111 Mean (SD) Age: 18.5 (SD 3.0) | Dance schools and performing productions in Ireland and other countries. | General Self- Efficacy Scale (GSES) | High psychological distress associated with injury with tension with people (47.3%), stress due to external factors (30.8%), general anxiety (29%) and performance anxiety (26.6%) reported most frequently by injured PIDs. No significant relationship found between the total number of injuries and psychological distress. |
| Davies & Mangion; 2002, Australia(13) | Cross-sectional | Professional instrumental musicians. N = 240; M: 135, F: 105. Mean (SD) Age:36.7(11). | Classical and non-classical music industry around Sydney, Australia | Bespoke survey of pain/symptoms, musical background, pain beliefs, workplace stress. | Musculoskeletal pain/symptoms significantly (<p=0.05) associated with high levels of playing-related muscle tension; high stress levels in conjunction with inadequate opportunities for rest/warmup; poor health and fewer years of playing experience. |
| Eliassen et al.; 2024, Norway(14) | Cross-sectional | Symphony orchestra musicians. N = 358. M:196, F:160. Unknown: 2. Mean (SD) Age: M: 47.7 (10.3); F: 43.4 (9.8). | Multiple professional orchestras in Norway | Hopkins Symptom Checklist-25 (HSCL-25). Bergen Insomnia Scale (BIS). | Higher experienced pain severity was associated with increased psychological distress (p <0.001) and sleep issues (p<0.001. |

| | | | | | |
|---|---|---|---|---|---|
| Hamilton et al.; 1989, United States(15) | Cross-sectional | Professional ballet dancers. N = 29; M;15, F:14. Mean (SD)Age: M:29.93(4.25); F: 29.23 (5.25) | New York City Ballet and American Ballet Theatre | Adult Personality Inventory (API). Occupational Environmental Stress scale (OES). Personal Strain Questionnaire (PSQ). Personal Resources Questionnaire (PRQ). | Dancers with greater numbers of injuries were significantly ($p<0.05$) more enterprising, dominant, extraverted, adjusted, and were more sociable but less withdrawn. Dancers with stress fractures were significantly ($p<0.05$) more dominant, extraverted, enterprising, assertive, and adjusted and their interpersonal styles were more sociable and less withdrawn than dancers without these problems. |
| Hilgenberg-Sydney et al.; 2020, Brazil(16) | Cross-sectional | Ballet dancers. N = 51; M:20; F:31 Mean (SD) Age: 31.5(12.6). | Guaira Theatre Dance School | State-Trait Anxiety Inventory (STAI). | No significant association between anxiety and the presence of TMDs. |
| Ioannou et al.; 2018, Germany(17) | Retrospective cohort with a follow-up questionnaire | Music students. N=186.M: 98. F: 88. Mean (SD) Age: 23 (2.7) | Outpatient clinic of the Institute of Music Physiology and Musicians Medicine | Competitive Trait Anxiety Inventory CTAI). State-Trait Anxiety Inventory (STAI). | Higher levels of self-doubt concern on the CTAI was significantly associated with poor motor performance ($p=0.005$). Higher levels of the trait anxiety were associated with elevated pain severity |
| Jabusch et al.; 2004, Germany(18) | Cross-sectional | N=70; M:37: F:33. Musicians with focal dystonia (FD): 20; Chronic pain (CP): 20; Healthy controls (HC): 30; Mean (SD)Age: FD 36.7 (6.4); CP 32.6 (8.7); HC 32.9 ( 5.4) | Outpatient clinic of the Institute of Music Physiology and Musician's Medicine in Hannover Germany | Revised version of the Freiburg Personality Inventory (FPI-R); Questionnaire for Competence and Control Orientations (QCC). Bespoke perfectionism and anxiety scale. | Anxiety disorders occurred more often in musicians with focal dystonia and chronic pain compared to healthy musicians ($p<0.05$ and $p<0.01$ respectively). Musicians with focal dystonia had higher perfectionistic tendencies than controls. Patients with chronic pain had more somatic complaints, and were more emotional compared to controls. |

| | | | | | |
|---|---|---|---|---|---|
| Junge & Hauschild;2023, Germany(19) | Cross-sectional | Professional dancers. N=147. M:65;F:82; Mean (SD)Age:27.1 ( 5.3) | Six German state theatres/opera houses | Patient Health Questionnaire (PHQ-9). Generalised Anxiety Disorder Assessment (GAD-7). Eating-Disorder-Examination-Questionnaire (EDE-QS). | For both genders, the PHQ-9 and GAD-7 sum scores were significantly related ($P<0.05$) to severity of musculoskeletal pain in the past seven days, |
| Kallusky et al.; 2023, Germany(20) | Longitudinal | Music students. N = 62 M: 15; F: 47. Mean (SD) Age: 22.8 (3.3) | Hannover University of Music, Drama and Media | Beck Depression Inventory II (BDI-II). Hospital Anxiety and Depression Scale (HADS. | Music students with pain had higher depression (BDI-II, $p < .05$) and anxiety (HADS, $p < .05$) scores. |
| Kaneko et al; 2005, Brazil(21) | Cross-sectional observational | Professional symphony orchestra musicians (N=241, M=168, F=73). Mean (SD) Age: 32.4 (10.6) | Six major symphony orchestras in São Paulo, Brazil | Beck Anxiety Inventory (BAI): Visual Analogue Scale (VAS): Emotional Stress; | Emotional stress & fear correlated strongly with longer pain durations. Sleep disorders also correlated ($p<0.001$). A statistically significant correlation was found between VAS score and the degree of emotional stress-related interference with the performance ($p = 0.014$) |
| Kaufman-Cohen & Ratzon; 2011, Israel(22) | Cross-sectional | Professional orchestral musicians. N=59. M:29. F:30. Mean (SD) Age: 42.9 (11.43). | Three major Israeli classical orchestras | National Institute for Occupational Safety and Health (NIOSH) Generic Job Stress Questionnaire | In univariate analysis, psychosocial job stress was somewhat associated with PRMDs, but final model found only biomechanical factors (instrument weight, daily hours) predicted. Concluded physical demands overshadow psychosocial stress in orchestral musician injuries. |

| | | | | | |
|---|---|---|---|---|---|
| Kenny & Ackermann; 2015, Australia(23) | Cross-sectional | Professional orchestral musicians. N=377: M:184. F:192. Mean (SD) Age: 42.1 (10.3). | Eight major Australian orchestras (including pit orchestras). | Kenny Music Performance Anxiety Inventory(K-MPAI).<br>Trait Questionnaire of the State-Trait Anxiety Inventory (STAI-T).<br>Social Phobia Inventory (SPIN).<br>PRIME-MD Patient Health Questionnaire (PRIME-MD PHQ) | Significant association between pain severity and depression (p=0.021)<br>Minimal effect of social anxiety. |
| Korte et al.; 2023, United Kingdom(24) | Cross-sectional (predictive model) | Music under/postgraduate vs. non-musicians (N=103). 58% female, Mean Age=~23.6. 62% with advanced classical training | University of Sheffield and allied colleges in United Kingdon | Örebro Musculoskeletal Pain Screening(Ö–MPSQ).<br>Hospital Anxiety and Depression Scale (HADS).<br>Cambridge Depersonalisation Scale (CD-9).<br>Copenhagen Burnout Inventory (CBI).<br>Brief COPE.<br>Pittsburgh Sleeping Quality Index(PSQI). | Found correlation between negative stress & longer durations of pain, but not clearly with catastrophising. Group testing showed that university non-musicians' pain catastrophising was significantly worse when compared to music college musicians. Music college musicians and university musicians were less prone to maladaptive pain processes, despite perceiving pain for significantly longer. |
| Lamontagne & Bélanger; 2015, Canada(25) | Cross-sectional pilot study | University-level orchestra music students N=59: M:23. F:36. Mean (SD) Age: 22(3). | Four Canadian university orchestras | Pain Anxiety Symptom Scale-Short (PASS-20).<br>Performance Anxiety Questionnaire (PAQ).<br>Positive and Negative Affect Scale (PANAS).<br>Minnesota Satisfaction Questionnaire-Short (MSQ). | Lower occupation satisfaction was found to be associate with the presence of pain (p=.02).<br>Performance anxiety was positively correlated with severity of pain (p=0.01) as was all pain-related anxiety (p<.001)<br>Performance anxiety and pain-related anxiety were found to explain 31% of the variance in pain severity (p<0.001)<br>No link with self-esteem or negative affect. |

| | | | | | |
|---|---|---|---|---|---|
| Leaver et al.; 2011, United Kingdom(26) | Cross-sectional survey | Professional orchestral musicians. N=243: M:136; F:107. Mean Age: 44. Range:23-64. | Six major British orchestras | Short-Form-36 -Mental health section (SF-36). Karasek Model: demand, support and control at work. Performance anxiety (instrument not specified). | Occupational psychosocial factors were associated somewhat inconsistently with regional pain, but ORs of $\geqslant 1.5$ were found in relation to low choice (for low-back and wrist/hand pain) and low support (for low-back, neck and shoulder pain), and an OR of 0.6 in relation to job insecurity and wrist/hand pain. |
| Liederbach & Compagno; 2001, United States(27) | Retrospective | Professional , university & clinic attending dancers. N = 644 . Professional: N=123: M:54. F:69: Mean (SD) Age: 24.6 (4.9). Student: N:282:M:90. F:192.Mean (SD) Age: 19.7 (2.2). (Clinic dancers not reported here as level not established.) | Professional ballet companies, outpatient clinics, universities | Eating Disorders Inventory 2 (EDI-2). Profile of Mood States (POMS). | Significant relationships between fatigue, overtraining, dieting behaviours, and increased injury risk. EDI-2 scores indicated higher perfectionism and body dissatisfaction among injured dancers ($p < .0001$). |
| Mainwaring et al.; 1993, Canada(28) | Prospective | Female university students majoring in dance. N = 39 F: 39. Mean (SD) Age: 21( 0.05) | University setting | Life Experiences Survey (LES). Dance Experiences Survey (DES). Rosenberg Self-Esteem Scale. | Negative stress significantly correlated with longer injury duration ($p < .05$). Positive stress inversely correlated with injury duration ($p < .05$). No significant relationship between self-esteem and injury duration. |

| | | | | | |
|---|---|---|---|---|---|
| Matei & Ginsborg;2020, United Kingdom(29) | Cross-sectional | Music students: N=111.M:47. F:64. Median: 22 (Range: 18-31). | Seven conservatoires in the United Kingdom. | Hospital Anxiety and Depression Scale (HADS). | Bodily pain interfering with practice and performance was associated with anxiety (HADS: rs = 0.438, CI = 0.089–0.562, p < 0.001).<br>Frequency of PRMDs was associated with anxiety (HADS: rs = 0.438, CI = 0.168–0.661, p < 0.001).<br>Severity of PRMDs was also associated with anxiety (HADS: rs = 0.340, CI = 0.078–0.572, p < 0.001). |
| Mathisen et al.; 2022, Norway(30) | Cross-sectional | Professional dance students. N = 124. M = 14, F = 110; Mean (SD)Age 20.7 ( 2.9) | Professional dance schools in Norway | Short Symptom Check List (SCL-10).<br>Resilience Scale for Adults (RSA).<br>Rosenberg Self-Esteem Scale (RSS).<br>Children and Adolescent Perfectionism Scale (CAPS).<br>Body Appreciation Scale-2 (BAS-2).<br>Eating Disorder Examination questionnaire (EDE-q) | In females, the authors evaluated whether training volume, symptoms of amenorrhea, academic year, BMI, BAS-2, EDE-q or age could explain the likelihood of injuries.<br>A significant model included BMI with OR = 1.39 (99% CI: 1.0, 1.9), academic year with OR = 2.86 (99% CI: 1.3, 6.2) and LEAF-q score with OR = 1.33 (99% CI: 1.1, 1.6) and had a specificity of 71% and sensitivity of 85%.<br>No equivalent model was reported for males/ |
| Michaels et al; 2023, United States(31) | Cross-sectional | Collegiate dancers. N = 198 M: 14; F: 184. Age: Not reported | University dance program | SCOFF Questionnaire.<br>National Athletic Trainers Association Mental Health Screening Tool (NATA-MH). | Symptoms of depression and anxiety associated with history of injury (p = .033) and active injuries (p = .039).<br>History of eating disorder associated with active injuries (p = .005). |

| | | | | | |
|---|---|---|---|---|---|
| Nordin-Bates et al.; 2011, United Kingdom.(32) | Cross-sectional | Ballet and contemporary dancers. N = 216 M: 17.4%; F: 82.6%. Mean (SD) Age: 21.6.(SD 5.6) | Dance college/university, conservatories, pre-professional, recreational/studio. | Rosenberg Self-Esteem Scale (RSES) | Injury is not related to dancers' self-esteem or at least not when injuries are mild or moderate, whether currently or previously injured. |
| Paris- Alemany et al.; 2022, Spain.(33) | Cross-sectional | Professional dancers. N = 34 M: 6; F: 28. Mean (SD) Age: 23.3 (3.8) | Higher conservatory of dance | Pain Catastrophizing Scale (PCS). Tampa Scale for Kinesiophobia (TSK). Fear-Avoidance Beliefs Questionnaire (FABQ). | Higher pain catastrophising and fear-avoidance beliefs associated with increased pain severity ($p < .05$). Fear of movement correlated with lower self-efficacy in managing pain ($p < .05$). Dancers with acute pain experienced worse psychological symptoms indicated by the fear of harm subscale of TSK-11 ($p = .04$; $t=-2.08$; $d=0.72$) |
| Patterson et al.; 1998, United States.(34) | Prospective | Ballet dancers. N = 46 M: 15; F: 31. Mean (SD) Age: 26.23 ( 4.14) | Professional ballet company | Perceived Events Scale (PES). Social Support Index (SSI). | High life stress and low social support significantly associated with injury ($p < .01$). Social support moderated the stress-injury relationship (interaction $p < .025$). |
| Pereira et al.; 2014, Brazil.(35) | Cross-sectional | Orchestra musicians. N = 22 M: 17; F: 5. Mean (SD) Age: 26.55 (11.33) | Southern Brazilian orchestra | Job Stress Scale (JSS). | High work demands associated with increased musculoskeletal complaints ($p < .05$). Social support perceived positively by all participants. High strain linked to most severe complaints ($p < .05$). |

| | | | | | |
|---|---|---|---|---|---|
| Ramel & Moritz; 1998, Sweden(36) | Cross-sectional | Professional ballet dancers. N = 64 M: 26; F: 38. Mean Age: 29 (Range: 17-47) | Royal Ballet in Stockholm | Bespoke questionnaire: psychosocial factors (job satisfaction, job influence, use of one's own capacity, social support, general stress and performance stress). | Muscular tension before performances associated with incapacitating pain (POR = 4.2, p = .03).<br>Dissatisfaction with work linked to higher prevalence of incapacitating pain (POR = 3.7, p = .03).<br>Not being able to use one's full capacity or experiencing increased tension on daily life increased the prevalence of incapacitation for work. "Capacity" and "general stress" did not contribute significantly. |
| Rip et al.; 2006, Canada.(37) | Cross-sectional | Dance students. N = 81 M: 7; F: 68. Unspecified: 6. Mean Age: 22.1 (Range: 15-31) | Department of Dance at University of Quebec, Montreal | Passion for Dance Scale (PDS).<br>Dancer Injury Profile Questionnaire (DIPQ) | Harmonious passion negatively associated with prolonged suffering from acute injuries (r = -.44, p = .03), positively associated with adaptive coping behavioUrs (p < .01) Obsessive passion linked to prolonged suffering from chronic injuries (r = .42, p = .03), rigid persistence (p < .05), and pride preventing treatment (r = .40, p = .000). |
| Rodríguez-Romero et al.; 2016, Spain.(38) | Cross-sectional | Music conservatory students. N = 206 M: 112; F: 94. Mean (SD) Age: 23.5 ( 4.1) | Higher Conservatories of Music in A Coruña and Vigo, Spain | SF-36, version 2: Mental Component Summary | Poor mental health associated with greater disability in neck (B = -0.3) and upper limbs (B = -0.4). |

| | | | | | |
|---|---|---|---|---|---|
| Sandell et al.; 2009, United States.(39) | Cross-sectional | Professional percussionists. N = 279 M: 236; F: 43. Mean (SD) Age: 28 ( 9.7) | University of North Texas survey. Online survey | University of North Texas Musician Health Survey (UNT-MHS). | Of those who reported one or more PRMDs, 76.9% also reported moderate to high stress levels, and of those who reported no PRMDs, 68.4% reported moderate to high stress levels (p = 0.193). Out of the whole sample of percussionists, the highest prevalence of stress-related health problems was reported for fatigue (49.5%), depression (35.1%), and stage fright (30.8%). |
| Scech.; Poland; 2021.(40) | Cross-sectional | Professional opera singers. N = 225. M:91 F:134. Age 18–67 years. | Six opera theatres in Poland | Neuroticism Ekstraversion Openness–Five Factor Inventory (NEO-FFI). Formal Characteristics of Behaviour–Temperament Inventory (FCB-TI). | Respondents with voice disorders were characterized by higher neuroticism i.e., the personality trait which increases susceptibility to stress (n = 129; p = 0.012) |
| Shrier I, Hallé M.; 2011; Canada.(41) | Historical cohort study | Circus artists. N=47 M: 30; F:17. Age: Data not provided | Cirque du Soleil training programme | Recovery-Stress Questionnaire for Athletes (REST-Q). | Self-efficacy Is associated with a > twofold (Risk ratio (RR): 2.6) increase in the risk of injury. Additionally, a high level of social stress (RR:1.9) or a low level of either success (RR: 1.5) or personal accomplishment(RR: 1.6) may also be predictive of injury. |
| Schwartz et al.; 2024, Hungary.(42) | Cross-sectional | Hungarian professional female dancers. N = 168 M: 0; F: 168. Mean (SD) Age: 32 (10.89) | Hungarian dance companies and online closed dance profile social media groups | Recovery-Stress Questionnaire for Athletes (RESTQ-Sport). Mental Health Test (MHT). | Correlations showed a statistically significant relationship between certain body part complaints and perceived stress. Pain correlated with stress in lower back (P = .01, r=.2), hip (P = .04, r=.14), and ankle (P = .04, r=.15). Resilience positively correlated with well-being (P < .001). |

| | | | | | |
|---|---|---|---|---|---|
| Skwiot et al.; 2014, Poland(43) | Cross-sectional | Athletes and dancers. N = 207. Dancers: 82: M: 18; F:64 . Mean (SD) Age: 21.5 (2.6) | University and professional settings | Sport Multidimensional Perfectionism Scale-2 (S-MPS-2).<br>Athlete Burnout Questionnaire (ABQ). | Athletes with high perfectionism and chronic pain reported the highest burnout (M = 2.37, SD = 0.68). Significant correlations between perfectionism and burnout components ($p < .05$). Pain moderated the relationship between perfectionism and burnout. |
| Smith et al.; 2000, United States.(44) | Prospective | Professional ballet dancers. N = 46 M: 15; F: 31. Mean (SD) Age: 26.23 ( 4.14) | Major ballet company in Western United States | Sport Anxiety Scale (SAS).<br>Perceived Events Scale (PES). | Stress experienced from major negative events was not a significant predictor of injury, nor did it interact with any of the SAS scales to predict injury variance. Cognitive and somatic anxiety moderated the relation between daily stress and injury ($p < .015$). There was a positive stress-injury correlation for the high anxiety dancers and a low negative correlation for the low anxiety dancers. High somatic anxiety correlated with increased injury time loss ($r = .41$, $p < .05$), as did worry ($r=0.36$), and concentration disruption ($r=.25$). Daily stress and somatic anxiety interaction explained 18% of injury variance. |
| Steemers et al.; 2020, Netherlands.(45) | Retrospective | Classical music students. N = 46 M: 18; F: 28. Mean (SD) Age: 21.3 ( 3.09) | Classical Music Department, Codarts Rotterdam, University of the Arts | Mental Health Inventory-5 (MHI-1). | Students with PRMDs scored lower on general health ($p = .012$). 45.7% had poor mental health (MHI-5 $\leqslant$ 60). No significant difference in mental health between students with and without PRMDs ($p=0.522$). |
| Steinmetz et al.; 2014, Germany(46) | Cross-sectional | Professional orchestral musicians. N = 408 M: 236; F: 172. Mean (SD) Age: 43.9 (10.3) | Classical orchestras in Berlin, Saxony, and Saxony-Anhalt | Bespoke questionnaire including stage fright self-assessment: Likert Scale - 0 (no stage fright) to 10 (worst imaginable stage fright). | Stage fright increased the occurrence of musculoskeletal pain between 1.4 and 2.9 times |

| | | | | | |
|---|---|---|---|---|---|
| Thomson et al.; 2020, United States(47) | Cross-sectional | Cohort of dancers N=185 and athletes: N=102. Dancers: M: 32; F: 105; Mean (SD) Age: 22.59(5.26) | Local University and surrounding community in Los Angeles. | Adverse Childhood Experiences (ACE).<br>Beck Anxiety Inventory (BAI-II).<br>Difficulties in Emotion Regulation Scale (DERS).<br>Dissociative Experience Scale—II (DES-II).<br>Traumatic Events Questionnaire (TEQ) | Participants (both dancers and athletes) who underwent orthopaedic surgery had more cumulative trauma and more difficulty with emotional regulation.<br>In both dancers and athletes, increased exposure to childhood and adult traumatic events were significant predictive factors associated with injury. |
| Topoglu et al.; 2018, Turkey(48) | Cross-sectional | Professional orchestral musicians. N = 220 M: 121; F: 99. Mean (SD) Age: 42.4 (11.3) | State symphony orchestras in Turkey | General Health Questionnaire-12 (GHQ-12).<br>Kenny Music Performance Anxiety Inventory (KMPAI) (Turkish version).<br>Bespoke health status questionnaire: Including performance anxiety and coping with performance anxiety. | The median KMPAI score of the musicians who had no musculoskeletal symptoms was 22 (range 7.0–46.0), and the median KMPAI score of the musicians who had at least one symptom was 34 (20.0–55.0). There was a significant difference between the two groups (U=683.5, *p*=0.04). |
| van Winden et al.; 2020, Netherlands(49) | Prospective | Contemporary dance students. N = 99 M: 28; F: 71. Mean (SD) Age: 19.2 (1.5) | Codarts Rotterdam University of the Arts | Athletic Coping Skills Inventory–28 (ACSI-28).<br>Perfectionism Inventory *Dance version) (PI).<br>Self-Regulation Questionnaire (short-form) (SSRQ). | No significant associations between independent variables and substantial injuries in follow up. However, a non-significant trend (p = 0.059) was visible for injury history , with coping skills (OR: 0.91; 95% CI: 0.84–0.98) identified amongst significant risk factors. |
| van Winden et al.; 2021, Netherlands(50) | Prospective | First-year contemporary dance students. N = 186 M: 59; F: 127. Mean (SD) Age: 19.21 (1.35) | Codarts Rotterdam University of the Arts | Bespoke visual analogue scale evaluating stress (VAS) 1-00. | Injured and substantially injured students reported higher stress scores than injury-free peers ($p < .05$). Stress levels increased before and during injury occurrence ($p = .051$ for all injuries; $p = .018$ for substantial injuries). |

| | | | | | |
|---|---|---|---|---|---|
| Zao et al.; 2024, Portugal(51) | Cross-sectional | Mixed cohort professional (N=291) and student musicians* (N= 294). Professional: M:109. F: 76. Mean (SD) Age:42.2 (11.9).<br><br>*Unable to distinguish <18 - all student cohort excluded from analysis. | Six academic & six professional music institutions. Randomly selected professional musicians. | 10-item Kessler Psychological Distress Scale.<br>Modified Fatigue Impact Scale (MFIS).<br>Frost Multidimensional Perfectionism Scale.<br>Short Form 36 Health Survey (SF-36). | Psychological distress (anxiety and depression: $p<0.001$) and perfectionism trait ($p=0.001$) among several psycho-socio-biodemographic and performance-related variables associated with the development of performance-related pain. |

BMI: Body mass index. F: Female. M: Male. MDE: Musculoskeletal disorder episodes; MHC: musculoskeletal health complaints; MPA: Music performance anxiety. OR: Odd's ratio. p: indicating statistical significance. PRMD: Performance-related musculoskeletal disorder. POR: Prevalence odd's ratio. PTSD: Post-traumatic stress disorder. r: Correlation coefficient. SD: Standard Deviation. TMD: Temporomandibular Disorder; TPP: Trigger point pain

**Supplementary Table 1: Details of included studies.**
