## Supplementary material for "Psychosocial risk factors for injury in performing artists: A scoping review of screening and predictive instruments": Supl 2: List of instruments

| **Instrument Name** | **Instrument Description** | **Primary Outcome Measure (s) Evaluated** | **Psychometric Testing in Dancers/Musicians/Vocalists/ Circus** | **Psychometric Properties** |
|---|---|---|---|---|
| Adult Personality Inventory (API)(1) | 324-item scale assessing normal-range adult personality traits across personal characteristics, interpersonal style, and career factors | Personality traits | Not reported | N/A |
| Adverse Childhood Experiences (ACE)(2) | 10-item questionnaire asking about experiences of abuse, neglect, and household challenges before the age of 18 | Childhood adversity mediated negative health outcomes in adulthood; e.g. risk of chronic diseases, mental health disorders, and premature mortality. | Not reported | N/A |
| Alcohol Use Disorders Identification Test (AUDIT)(3) | 10-item questionnaire that screens for hazardous or harmful alcohol consumption | Alcohol addiction | Not reported | N/A |
| Anxiety and Depression Detector (ADD)(4) | 5-item brief screening tool designed to identify individuals at risk for anxiety and depressive disorders in primary care settings | Anxiety and/or depression | Not reported | N/A |

| | | | | |
|---|---|---|---|---|
| Athletic Coping Skills Inventory (ACSI-28)(5) | A self-report assessment designed to evaluate athletes' psychological coping skills across seven key areas including: Coping with adversity, peaking under pressure,<br>goal setting and mental preparation,<br>concentration,<br>freedom from worry<br>confidence and achievement motivation<br>coachability | Psychological coping skills | | |
| Athlete Burnout Questionnaire (ABQ)(6) | 15-item self-report tool that assesses burnout in athletes across three dimensions: emotional and physical exhaustion, reduced sense of accomplishment, and devaluation of sport participation | Burnout | Tested for validity in dancers.(7)<br><br>Not evaluated in musicians, vocalists or circus performers. | A confirmatory factor analysis was conducted on the modified 15-item ABQ for dancers and athletes, which did not fit the model fit criteria. |
| Beck Anxiety Inventory (BAI-II)(8) | 21-item self-report questionnaire | Anxiety | Not reported | N/A |

| | | | | |
|---|---|---|---|---|
| | designed to assess the severity of anxiety symptoms in adults and adolescents. | | | |
| Beck Depression Inventory II (BDI-II)(9) | 21-item self-report inventory measuring the severity of depression in adolescents and adults | Depression | Evaluated for internal consistency in dancers.(10)<br><br>Not evaluated in musicians, vocalists or circus performers. | The study reported high internal consistency, with Cronbach's alpha coefficients of 0.86 for pretreatment, 0.91 for posttreatment, and 0.91 for follow-up measurements. |
| Bergen Insomnia Scale(11) | 6-item self-report questionnaire designed to assess insomnia symptoms and is rated based on frequency over the past month. | Sleep disturbance | Not reported | N/A |
| Body Appreciation Scale-2 (BAS-2)(12) | 13-item self-report questionnaire that assess body appreciation and positive body image. | Body image | Internal consistency tested in recreational pole-dancers.(13)<br><br>Not evaluated in musicians, vocalists or circus performers. | High internal consistency was reported with Cronbach's α values for BAS-2 pre-training and post-training scores were 0.95 |

| | | | | |
|---|---|---|---|---|
| | | | | and 0.94, respectively |
| Brief COPE(14) | A self-report questionnaire that assesses a range of coping strategies individuals use in response to stress, consisting of 28 items across 14 coping subscales. | Coping with stress | A modified COPE for ballet dancers was evaluated for reliability (internal consistency), factor structure and construct validity.(15)<br><br>Not evaluated in musicians, vocalists or circus performers. | Cronbach's alpha values for different subscales ranged from 0.60 to 0.85, indicating acceptable to good reliability for most coping strategies.<br><br>Factor analyses supported the scale's ability to differentiate between coping strategies.<br><br>The scale demonstrated good construct validity in identifying coping strategies linked to competitive anxiety and stress among dancers. |
| Cambridge Depersonalisation Scale (CD-9)(16) | 29-item self-administered questionnaire that measures the severity | Depersonalisation | Not reported | N/A |

| | | | | |
|---|---|---|---|---|
| | of depersonalisation experiences, such as feeling detached from oneself or observing one's thoughts and actions from outside the body. | | | |
| Children and Adolescent Perfectionism Scale (CAPS)(17) | 22-item self-administered questionnaire measuring perfectionistic tendencies in young people, including aspects such as self-criticism, concern over mistakes, and the pursuit of high standards. | Perfectionism | Internal consistency tested in competitive Irish dancers.(18)<br><br>Not evaluated in musicians, vocalists or circus performers. | CAPS demonstrated satisfactory internal consistency, with Cronbach's alpha coefficients ranging from 0.70 to 0.84 |
| Cohen Perceived Stress Scale(19) | 10-item self-administered questionnaire measuring the degree to which individuals perceive their life as stressful, assessing feelings of unpredictability, uncontrollability, and overload in daily life | Stress | Internal consistency and construct validity tested in Indian Kathak dancers.(20)<br><br>Not evaluated in musicians, vocalists or circus performers. | Internal consistency: Cronbach's alpha coefficients of 0.76 for dancers. The scale's construct validity was also confirmed. |

| | | | | |
|---|---|---|---|---|
| Competitive Trait Anxiety Inventory (CTAI)(21) | 15-item self-administered questionnaire that measures an individual's level of trait anxiety in competitive situations, assessing the tendency to experience anxiety in competitive settings. | Competition anxiety | Internal consistency tested in 20 musicians with focal dystonia.(22)<br><br>Not evaluated in dancers, vocalists or circus performers. | Cronbach's alpha values of 0.81 for the "somatic anxiety" subscale and 0.83 for the "self-doubt concern" subscale, indicating satisfactory internal consistency. |
| Copenhagen Burnout Inventory (CBI)(23) | 19-item self-administered questionnaire measuring burnout across three dimensions: personal burnout, work-related burnout, and client-related burnout, assessing physical and psychological exhaustion | Burnout | Not reported | N/A |
| Coping Inventory for Stressful Situations (CISS)(24) | 48-item self-administered measuring three coping styles—task-oriented, emotion-oriented, and avoidance-oriented— | Coping with stress | Internal consistency tested in a cohort of pre-professional and professional dancers.(25)<br><br>Not evaluated in musicians, vocalists or circus performers. | Excellent internal consistency with Cronbach's alpha coefficients of 0.84 for task-oriented focus, 0.90 for emotion-oriented |

| | | | | |
|---|---|---|---|---|
| | assessing how individuals respond to stress. | | | focus, and 0.86 for avoidance-oriented coping. |
| Coping Strategies Questionnaire (CSQ)(26) | 50-item self-administered tool measuring cognitive and behavioural coping strategies used to manage pain, including catastrophising, ignoring sensations, reinterpreting pain, and seeking social support. | Coping | Not reported | N/A |
| Core Self Evaluation Scale (CSE)(27) | 12-item self-administered questionnaire measuring an individual's fundamental self-assessments, including self-esteem, generalized self-efficacy, locus of control, and emotional stability (neuroticism) | Core self-evaluation | Not reported | N/A |

| | | | | |
|---|---|---|---|---|
| Dancer Injury Profile Questionnaire(28) | 21 items assessing physical injury characteristics as well as coping behaviours in responses to pain and injury, items assessing health-promoting, as well as health-undermining coping behaviours. | Coping and health promotion behaviours. | Not reported | N/A |
| Depression Anxiety Stress Scale (DASS)(29) | 42-item self-report instrument designed to measure three related negative emotional states, namely depression, anxiety and stress. | Depression, anxiety and stress. | Not reported | N/A |
| Difficulties in Emotion Regulation Scale (DERS)(30) | 36-item self-report measure of six facets of emotion regulation, namely Nonacceptance of Emotional Responses, Difficulties in Engaging in Goal-Directed Behaviour, Impulse Control | Emotional regulation | Not reported | N/A |

| | | | | |
|---|---|---|---|---|
| | Difficulties, Lack of Emotional Awareness, Limited Access to Emotion Regulation Strategies, and Lack of Emotional Clarity | | | |
| Dissociative Experience Scale—II (DES-II)(31) | 28-item self-administered questionnaire related to experiences of dissociation, such as depersonalization, derealization, amnesia, and absorption. | Dissociation | Not reported | N/A |
| Eating Disorders Inventory 2 (EDI-2)(32) | 91-items self-administered questionnaire of subscales assessing attitudes, behaviours, and psychological traits related to eating disorders, such as drive for thinness, bulimia, and body dissatisfaction. | Disordered eating. | Not reported | N/A |
| Eating disorder examination | 28-items self-administered questionnaire that | Disordered eating. | Not reported | N/A |

| | | | | |
|---|---|---|---|---|
| questionnaire (EDE-QS)(33) | assess the frequency and severity of eating disorder behaviours and attitudes, such as restraint, eating, shape, and weight concerns. | | | |
| Fear-Avoidance Beliefs Questionnaire (FABQ)(34) | 16-items self-administered questionnaire including two subscales that assess fear-avoidance beliefs related to physical activity and work in individuals with back pain or musculoskeletal disorders. | Fear-avoidance beliefs | Not reported | N/A |
| Fonseca Anamnestic Questionnaire(35) | 10-item self-administered questionnaire designed to assess symptoms related to temporomandibular disorders (TMD), such as pain, jaw movement, and headaches. | Tension (and other physical factors) related to TMD | Not reported | N/A |

| | | | | |
|---|---|---|---|---|
| Formal Characteristics of Behaviour–Temperament Inventory (FCB-TI)(36) | 120 yes/no questions, divided into six subscales measuring traits such as: briskness, perseverance, sensory sensitivity, emotional reactivity, endurance, and activity | Temperament | Not reported | N/A |
| Freiburg Personality Inventory - Revised (FPI-R)(37) | 138 item self-administered questionnaire assessing various personality traits, such as emotional stability, social behaviour, and self-confidence | Personality | Not reported | N/A |
| Frost Multidimensional Perfectionism Scale(38) | 35-item question self-report measure with four sub-scales of perfectionism. | Perfectionism | Not reported | Note: Abridged version of this instrument has been found to have internal reliability in elite vocalists. Cronbach's alpha = 084.(39) |
| General Health Questionnaire-12(40) | 12-items self-administered questionnaire to | Psychological well-being. | Not reported | N/A |

| | | | | |
|---|---|---|---|---|
| | assess psychological well-being, focusing on symptoms of anxiety, depression, and social dysfunction | | | |
| General Self-Efficacy Scale (GSES)(41) | 10-item self-administered questionnaire which measures an individual's belief in their ability to cope with a variety of challenging or stressful situations | Self-efficacy/ coping | Not reported | N/A |
| Generalised Anxiety Disorder Assessment (GAD-7)(42) | 7-item self-reported instrument that is used to measure or assess the severity of generalised anxiety over the previous two weeks. | Anxiety | Internal consistency tested in Indian Kathak dancers.(20)<br><br>Not evaluated in musicians, vocalists or circus performers. | Cronbach’s α for the internal consistency of the GAD-7 was 0.84 for Kathak dancers |
| Hopkins Symptom Checklist-25 (HSCL-25)(43) | 58-item checklist scored on five underlying symptom dimensions—somatization, obsessive-compulsive, interpersonal | Anxiety, depression, interpersonal sensitivity. | Internal consistency tested in cohort of professional Norwegian musicians.(44)<br><br>Not evaluated in dancers, vocalists or circus performers. | Cronbach's α (internal consistency) of 0.93 for the total HSCL-25 score, indicating strong reliability. |

| | | | | |
|---|---|---|---|---|
| | sensitivity, anxiety and depression. | | | |
| Hospital Anxiety and Depression Scale (HADS)(45) | 14-item self-assessed tool that measures anxiety and depression levels in individuals, with 7 items for each dimension. | Anxiety and depression | Internal consistency tested in cohort of professional Danish musicians.(46)<br><br>Not evaluated in dancers, vocalists or circus performers. | Cronbach's α (internal consistency) of 0.89 for HADS-Anxiety, and 0.82 for HADS-Depression scores. |
| Job Stress Scale (JSS)(47) | 13 item self-administered scale that measures job stress, focusing on various factors like workload, role conflict, and social support | Job stress | Not reported | N/A |
| Karasek Model: demand, support and control at work(48) | 49-item self-assessment questionnaire that measures job demands, decision latitude (control), and social support to evaluate workplace stress and its impact on employee well-being. | Occupational stress. | Not reported | N/A |
| Kenny Music Performance Anxiety | 40-item inventory that assesses an emotion- | Performance anxiety | Widely examined in multiple cohorts of musicians.(49) | K-MPAI consistently demonstrates a |

| | | | | |
|---|---|---|---|---|
| Inventory (K-MPAI)(49) | based theory of anxiety as it applies to anxiety in the context of music performance. | | Additionally evaluated in a cohort of 32 elite operatic chorus artists.(39)<br><br>Not evaluated in dancers or circus performers. | stable factorial structure, robust reliability, and strong utility for diagnostic purposes among musicians, across various studies and cultural contexts.(49)<br><br>Internal reliability of K-MPAI in vocalists: Cronbach's alpha = 0.944(39) |
| Kessler Psychological Distress Scale(50) | 10-item self-report questionnaire designed to measure the level of psychological distress based on anxiety and depressive symptoms experienced over the past 30 days. | Anxiety and Depression | Not reported | N/A |
| Life Experiences Survey (LES)(51) | 57-item self-report questionnaire which assess various life events and their impact, categorising | Life stressors | Not reported | N/A |

| | | | | |
|---|---|---|---|---|
| | them as either positive or negative stressors | | | |
| Mental Health Inventory-5(52) | 5-item self-administered questionnaire assessing psychological well-being and distress, including aspects like anxiety, depression, positive affect, and general well-being | Psychological well-being | Not reported | N/A |
| Mental Health Test (MHT)(53) | 17-item self-administered scale that assesses various aspects of mental health, including symptoms related to mood, anxiety, and general psychological well-being. | Mental health | Not reported | N/A |
| Minnesota Satisfaction Questionnaire-Short (MSQ)(54) | 100-item self-administered questionnaire assessing 20 different facets of job satisfaction. | Job satisfaction | Not reported | N/A |

| | | | | |
|---|---|---|---|---|
| Modified Fatigue Impact Scale (MFIS)(55) | 21-item self-administered questionnaire assessing fatigue on a person's physical, cognitive, and psychosocial functioning. | Multifactorial fatigue | Not reported | N/A |
| Musculoskeletal Pain Intensity and Interference Questionnaire (MPIIIQ)(56) | 9-item self-administered questionnaire that assesses the intensity of musculoskeletal pain and its impact on daily activities, mood, and performance in professional musicians. | Pain-related mood, performance. | Evaluated in professional musicians.(56)<br><br>Not evaluated in dancers, vocalists or circus performers. | Internal consistency: Cronbach's alpha of 0.91 for pain intensity and pain interference subscales. Test-Retest Reliability: intraclass correlation coefficients ranging from 0.78 to 0.82 and moderate to substantial reliability for pain interference items (coefficients ranging from 0.56 to 0.76). |

| | | | | |
|---|---|---|---|---|
| | | | | Factor Structure: Exploratory factor analysis revealed a stable two-factor structure, accounting for 71.3% of the variance, identifying distinct pain intensity and pain interference components. |
| National Athletic Trainers Association Mental Health Screening Tool(57) | 9-item self-report measure that asks for yes/no responses on a range of mental health topics. | General mental health. | Not reported | N/A |
| National Institute for Occupational Safety and Health (NIOSH) Generic Job Stress Survey(58) | Measures 13 different job stressors as well as a host of measures of individual distress and modifiers of the stress response | Job stress | Not reported | N/A |
| Neuroticism Ekstraversion Openness–Five Factor Inventory (NEO-FFI)(59) | 60 self-reporting statements rated on a 5-point scale measuring: neuroticism, extraversion, | Personality and adaptation capability | Not reported | N/A |

| | | | | |
|---|---|---|---|---|
| | openness to experience, agreeableness, and conscientiousness | | | |
| Occupational Environmental Stress (OES) scale(60) | 48 item scale which assess different aspects of stress in the workplace, focusing on both environmental and organizational factors that contribute to job stress. | Occupational stress | Not reported | N/A |
| Örebro Musculoskeletal Pain Screening Questionnaire (OMPSQ)(61) | 25-items self-completed tool assessing psychosocial and physical factors related to musculoskeletal pain, including pain intensity, disability, and the risk of developing chronic pain or disability. | Biopsychosocial factors associated with pain. | Not reported | N/A |
| Pain Anxiety Symptom Scale-Short (PASS-20)(62) | 20-item self-completed questionnaire that assesses the severity of pain-related | Fear avoidance, pain hypervigilance. | Not reported | N/A |

| | | | | |
|---|---|---|---|---|
| | anxiety by measuring symptoms such as fear, avoidance, and hypervigilance in response to pain. | | | |
| Pain Catastrophizing Scale (PCS)(63) | 13-item self-report measure of catastrophising in the context of actual or anticipated pain | Catastrophising | Not reported | N/A |
| Passion for Dance Scale (PDS)(28) | Composed of two six-item subscales assessing harmonious and obsessive passion for dance, as well as four passion criterion items measuring whether dance could be considered a "passion" for each participant | Passion for dance | Internal consistency tested in a cohort of mixed-genre dancers.(28)<br><br>Not evaluated in musicians, vocalists or circus performers. | Internal consistency indices of .78 and .84 were obtained for the harmonious and obsessive passion subscales, respectively. |
| Passion Scale (PS)(64) | Composed of two seven-item subscales assessing harmonious and obsessive passion for a given activity. | Passion | Not reported | N/A |

| | | | | |
|---|---|---|---|---|
| Patient Health Questionnaire-9 (PHQ-9)(65) | 9-item self-reported tool that assess the severity of depressive symptoms over the past two weeks, focusing on mood, interest, energy, and related aspects of depression | Mood, depression. | Not reported | N/A |
| Perceived Events Scale (PES)(66) | 207 questions pertaining to events covering a wide range of life domains. For each item, the participants indicated whether the event had occurred in the past 6 months. | Positive and negative life events. | Not reported | N/A |
| Perfectionism Inventory (Dance version)(67) | Includes seven scales assessing various domains of perfectionism.<br>The first three subscales constitute the factor conscientious perfectionism, while the latter four address self-evaluative perfectionism. | Perfectionism | Internal consistency evaluated in cohort of contemporary and ballet dancers.(67)<br><br>Not evaluated in musicians, vocalists or circus performers. | Internal reliability per subscale (Cronbach's alpha values) ranging from<br>0.74 to 0.89 |

| | | | | |
|---|---|---|---|---|
| Performance Anxiety Questionnaire (PAQ)(68) | Measures both cognitive anxiety (10 items) and physiological anxiety (10 items) related to musical performance in three different contexts (solo performance, chamber music performance and orchestral performance). | Performance anxiety. | Internal consistency evaluated in cohort of music students.(69)<br><br>Not evaluated in dancers, vocalists or circus performers. | Internal consistency of Cronbach's α = .90 and correlated at r = .60, p < .001 with the negative affect scale. |
| Personal Resources Questionnaire (PRQ)(70) | A 2-part measure of the multidimensional characteristics of social support. Part1 pertains to resources and supports, and Part 2 relates to self-help. | Situational and perceived social support | Not reported | N/A |
| Personal Strain Questionnaire (PSQ)(60) | 40-item self report instrument including four areas namely: Vocational Strain, Psychological Strain, Interpersonal Strain , Physical Strain. | Multidimensional strain | Not reported | N/A |

| | | | | |
|---|---|---|---|---|
| Pittsburgh Sleep Quality Index (PSQI)(71) | A self-report questionnaire that assesses sleep quality over a 1-month time interval. The measure consists of 19 individual items, creating 7 components that produce one global score | Sleep quality | Not reported | N/A |
| Positive and Negative Affect Scale (PANAS)(72) | This brief scale is comprised of 20 items, with 10 items measuring positive affect (e.g., excited, inspired) and 10 items measuring negative affect (e.g., upset, afraid). | Mood/emotion | Not reported | N/A |
| PRIME-MD Patient Health Questionnaire (PRIME-MD PHQ)(65) | A self-administered 1-page questionnaire consisting of 26 yes/no questions about the presence of symptoms and signs of common mental disorders during the past month | Mental disorder | Not reported | N/A |

| | | | | |
|---|---|---|---|---|
| Profile of Mood States (POMS)(73) | POMS measures six different dimensions of mood swings over a period of time. These include: Tension or Anxiety, Anger or Hostility, Vigor or Activity, Fatigue or Inertia, Depression or Dejection, Confusion or Bewilderment. | Mood | Not reported | N/A |
| Questionnaire for Competence and Control Orientations (QCC)(74) | QCC investigates features such as self-concept of abilities, internal control orientation, others control orientation, chance control orientation | Competence and control | Not reported | N/A |
| Recovery-Stress Questionnaire for Athletes (RESTQ-Sport)(75) | 52-item subjective questionnaire sensitive to the stress and recovery incurred by sports and general lifestyles | Stress and recovery | Dance version of the REST-Q has been developed and evaluated.(76)<br><br>Not evaluated in musicians, vocalists or circus performers. | Factor Structure: The RESTQ-Dance comprises 63 items organized into three factors:<br><br>Internal consistency: General Stress: 26 items (Cronbach's α = 0.92) |

| | | | | |
|---|---|---|---|---|
| | | | | Recovery: 27 items (Cronbach's α = 0.91)<br>Specific Stress: 10 items (Cronbach's α = 0.79)<br>Internal Consistency: Cronbach's alpha values for the factors indicate good internal consistency, with values ranging from 0.79 to 0.92. |
| Resilience Scale for Adults(77) | 25-item self-report scale designed to assess the level of resilience in adults. Includes dimensions of personal resilience, such as personal competence, acceptance of self and life, and social competence | Resilience and coping | Not reported | N/A |
| Rosenberg Self-Esteem Scale(78) | 10-item self-report scale that measures global self-worth by measuring both | Self-esteem | Not reported | N/A |

| | | | | |
|---|---|---|---|---|
| | positive and negative feelings about the self | | | |
| Short-form Self-Regulation Questionnaire(79) | 31-item instrument designed to assess their capacity for self-regulation; that is, the ability to plan, guide, and monitor behaviours in the face of changing circumstances | Self-regulation | Not reported | N/A |
| Short Symptom Check List (SCL-10)(80) | 10 questions scored from 0 to 4, and all are averaged into a global score, in which higher scores present higher severity of symptoms. A cut-off score of 1.85 indicates symptoms of mental health issues. | General health issues including mental health. | Not reported | N/A |
| Short-Form Health Survey SF36(81) | 36-item questionnaire that assesses health-related quality of life across eight domains, including physical functioning, pain, | Health related quality of life | Not reported | N/A |

| | | | | |
|---|---|---|---|---|
| | emotional well-being, and social functioning | | | |
| Sick, Control, One stone, Fat, Food (SCOFF) questionnaire(82) | A brief, 5-item measure that has been used to screen for anorexia nervosa and bulimia nervosa | Disordered eating | Not reported | N/A |
| Social Phobia Inventory (SPIN)(83) | 17-item self-rating scale for social anxiety disorder (social phobia). The scale is rated over the past week and includes items assessing each of the symptom domains of social anxiety disorder (fear, avoidance, and physiologic arousal) | Social phobia/ anxiety. | Not reported | N/A |
| Social Support Appraisals Scale (SS-A)(84) | 23-item scale measures perceived social support from family, friends, and other members of the immediate community. | Social support | Not reported | N/A |

| | | | | |
|---|---|---|---|---|
| Social Support Index(85) | Scale includes completed measures of the amount and quality of social support available to them from 20 different individuals (e.g., mother, father, coach, and best friend) and groups (e.g., their teammates and clubs or religious groups to which they belonged). | Social support | Not reported | N/A |
| Sport Anxiety Scale (SAS)(86) | 21-item measure of trait anxiety. It has separate subscales for somatic anxiety, and for two varieties of cognitive anxiety: worry, and concentration disruption. | Performance anxiety | Internal consistency reported in a cohort of professional ballet dancers.(86)<br><br>Not evaluated in musicians, vocalists or circus performers. | Internal consistency: Cronbach's α coefficients were 0.84 for somatic anxiety, 0.79 for Worry, and 0.76 for concentration disruption. |
| Sport Multidimensional Perfectionism Scale-2 (S-MPS-2)(87) | Six domains for evaluating the level of perfectionism regarding :personal standards ; concern over mistakes; | Perfectionism | Not reported | N/A |

| | | | | |
|---|---|---|---|---|
| | doubts about action; perceived parental pressure; perceived coach pressure; and need for organisation | | | |
| Stress and Coping Inventory (SCI)(88) | A self-report tool used to determine the current stress load and stress symptoms and to illustrate how to deal with stress using five coping strategies. It comprises 10 scales with 54 items. | Coping with stress | Not reported | N/A |
| State-Trait Anxiety Inventory (STAI-T)(89) | 20-item scale used to measure state (20 items) and trait (20 items) anxiety. | Anxiety | Not reported | N/A |
| Tampa Scale for Kinesiophobia-11 (TSK-11) (Spanish version)(90) | TSK-11 consists of two subscales, one related to fear of activity and the other related to fear of harm. The final score can range between 11 and 44 points, with higher scores indicating | Fear of activity/harm | Not reported | N/A |

| | | | | |
|---|---|---|---|---|
| | greater perceived kinesiophobia. | | | |
| Traumatic Events Questionnaire (TEQ)(91) | A self-report 11-item dichotomously scored instrument that assesses exposure to 9 different traumatic events | Legacy of trauma | Test-retest reliability reported in a cohort of undergraduate student dancers.(92)<br><br>Not evaluated in musicians/circus performers. | Test-retest of TEQ was found to be highly stable over this time span, with a correlation of .95 (p < .001) between the two TEQ scores. |
| University of North Texas Musician Health Survey(93) | A comprehensive self-report questionnaire designed to assess physical and mental health issues, performance-related injuries, and wellness behaviours among musicians. | Holistic wellness | Not reported | N/A |

**Supplementary Table 2: Instrument details**
