## Supplementary material for "Psychosocial risk factors for injury in performing artists: A scoping review of screening and predictive instruments": Supl 3: Search strings

**Initial search strategy: Medline**

("psychosocial factors"[Title/Abstract] OR "mental health"[Title/Abstract] OR "anxiety"[Title/Abstract] OR "depression"[Title/Abstract] OR "catastrophizing"[Title/Abstract] OR "catastrophising"[Title/Abstract] OR "psychological stress"[Title/Abstract] OR "burnout"[Title/Abstract] OR "cognitive factors"[Title/Abstract] OR "stress"[Title/Abstract])

AND

("pain"[Title/Abstract] OR "injury"[Title/Abstract] OR "musculoskeletal pain"[Title/Abstract] OR "chronic pain"[Title/Abstract] OR "musculoskeletal disorders"[Title/Abstract] OR "injury prevention"[Title/Abstract] OR "injury risk"[Title/Abstract] OR "workplace injury"[Title/Abstract])

AND

("performing artists"[Title/Abstract] OR "danc*"[Title/Abstract] OR "musician*"[Title/Abstract] OR "instrumentalist*"[Title/Abstract] OR singer*[Title/Abstract] OR vocalist*[Title/Abstract] OR actor*[Title/Abstract] OR "circus performer*"[Title/Abstract] OR "circus arts"[Title/Abstract] OR "performing arts"[Title/Abstract] OR "occupational performer*"[Title/Abstract]

AND "elite*"[Title/Abstract] OR "professional*"[Title/Abstract] OR "pre-professional *"[Title/Abstract] OR "non-recreational"[Title/Abstract])

AND

("screening tools" OR "predictor tools" OR "predictive tools" OR "questionnaire*" OR "outcome measures" OR "risk assessment tools" OR "assessment tools" OR "evaluation tools" OR "diagnostic tools" OR "measurement tools")

**Web of Science**

("psychosocial factors" OR "mental health" OR "anxiety" OR "depression" OR "catastrophizing" OR "catastrophising"OR "psychological stress" OR "burnout" OR "cognitive factors" OR "stress") (Abstract) and ("pain" OR "injury" OR "musculoskeletal pain" OR "chronic pain" OR "musculoskeletal disorders" OR "injury prevention" OR "injury risk" OR "workplace injury") (Abstract) and ("performing artists" OR "danc*" OR "musician*" OR "instrumentalist*" OR singer* OR vocalist* OR actor* OR "circus performer*" OR "circus arts" OR "performing arts" OR "occupational performer*" AND "elite" OR "professional*" OR "full-time student" OR "pre-professional*" OR "non-recreational performer*") (Abstract) and ("screening tools" OR "predictor tools" OR "predictive tools" OR "questionnaire*" OR "outcome measures" OR "risk assessment tools" OR "assessment tools" OR "evaluation tools" OR "diagnostic tools" OR "measurement tools") (All Fields) not ("CHILD*" OR "teen*" OR "ADOLESCEN*" OR "PEDIATRIC" OR "PAEDIATRIC") (Abstract)

**EMBASE**

('psychosocial factors' OR 'mental health' OR 'anxiety' OR 'depression' OR 'catastrophizing' OR 'catastrophising' OR 'psychological stress' OR 'burnout' OR 'cognitive factors' OR 'stress' OR 'mental' OR 'emotional') AND ('pain' OR 'injury' OR 'musculoskeletal pain' OR 'chronic pain' OR 'musculoskeletal disorders' OR 'injury prevention' OR 'injury risk' OR 'wound' OR 'workplace injury') AND ('perform* artist' OR 'danc*' OR 'musician*' OR 'instrumentalist*' OR 'singer*' OR 'vocalist*' OR 'theatre*' OR 'actor*' OR 'actress*' OR 'circus performer*' OR 'circus arts' OR 'performing arts' OR 'occupational performer*') AND ('elite' OR 'professional*' OR 'full-time student' OR 'pre-professional*' OR 'non-recreational performer*') AND ('screening' OR 'predictor' OR 'predictive tools' OR 'questionnaire*' OR 'survey*' OR 'outcome measures' OR 'risk assessment tools' OR 'assessment tools' OR 'assessment' OR 'inventory' OR 'evaluation tools' OR 'diagnostic tools' OR 'measur* tools') AND [adult]/lim AND 'human'/de

**Cochrane**

 'psychosocial factors' OR 'mental health' OR 'anxiety' OR 'depression' OR 'catastrophizing' OR 'catastrophising' OR 'psychological stress' OR 'burnout' OR 'cognitive factors' OR 'stress' OR 'mental' OR 'emotional' in Title Abstract Keyword AND 'pain' OR 'injury' OR 'musculoskeletal pain' OR 'chronic pain' OR 'musculoskeletal disorders' OR 'injury prevention' OR 'injury risk' OR 'wound' OR 'workplace injury' in Title Abstract Keyword AND ('perform* artist' OR 'dancing' OR 'dancer' OR 'musician*' OR 'instrumentalist*' OR 'singer*' OR 'vocalist*' OR 'theatre*' OR 'actor*' OR 'actress*' OR 'circus performer*' OR 'circus arts' OR 'performing arts' OR 'occupational performer*') in Title Abstract Keyword AND ('elite' OR 'professional*' OR 'full-time student' OR 'pre-professional*' OR 'non-recreational performer*') in All Text AND ('screening' OR 'predictor' OR 'predictive tools' OR 'questionnaire*' OR 'survey*' OR 'outcome measures' OR 'risk assessment tools' OR 'assessment tools' OR 'assessment' OR 'inventory' OR 'evaluation tools' OR 'diagnostic tools' OR 'measur* tools') in All Text - (Word variations have been searched)

**EBSCO: (Medline, Cinahl, Sports Discus, PsychInfo)**

Search Alert: "AB ( 'psychosocial factors' OR 'mental health' OR 'anxiety' OR 'depression' OR 'catastrophizing' OR 'catastrophising' OR 'psychological stress' OR 'burnout' OR 'cognitive factors' OR 'stress' OR 'mental' OR 'emotional' ) AND AB ( 'pain' OR 'injury' OR 'musculoskeletal pain' OR 'chronic pain' OR 'musculoskeletal disorders' OR 'injury prevention' OR 'injury risk' OR 'wound' OR 'workplace injury' ) AND AB ( 'perform* artist' OR 'danc* OR 'musician' OR 'instrumentalist' OR 'singer' OR 'vocalist' OR theater OR 'theatre*' OR 'actor*' OR 'actress*' OR 'circus performer*' OR 'circus arts' OR 'performing arts' OR 'occupational performer' ) AND TX ( 'elite' OR 'professional*' OR 'full-time student' OR 'pre-professional' OR 'non-recreational performer' ) AND TX ( 'screening' OR 'predictor' OR 'predictive tools' OR 'questionnaire*' OR 'survey*' OR 'outcome measures' OR 'risk assessment tools' OR 'assessment tools' OR 'assessment' OR 'inventory' OR 'evaluation' OR 'diagnostic' OR 'measur* tools' ) NOT ( children or adolescents or youth or child or teenager ) Apply equivalent subjects on 2024-11-14 02:34 PM"

**PubMed**

(((('psychosocial factors'[Title/Abstract] OR 'mental health'[Title/Abstract] OR 'anxiety'[Title/Abstract] OR 'depression'[Title/Abstract] OR 'catastrophizing'[Title/Abstract] OR 'catastrophising'[Title/Abstract] OR 'psychological stress'[Title/Abstract] OR 'burnout'[Title/Abstract] OR 'cognitive factors'[Title/Abstract] OR 'stress'[Title/Abstract] OR 'mental'[Title/Abstract] OR 'emotional'[Title/Abstract]) AND ('pain'[Title/Abstract] OR 'injury'[Title/Abstract] OR 'musculoskeletal pain'[Title/Abstract] OR 'chronic pain'[Title/Abstract] OR 'musculoskeletal disorders'[Title/Abstract] OR 'injury prevention'[Title/Abstract] OR 'injury risk'[Title/Abstract] OR 'wound'[Title/Abstract] OR 'workplace injury'[Title/Abstract])) AND ('perform* artist'[Title/Abstract] OR 'danc*'[Title/Abstract] OR 'musician'[Title/Abstract] OR 'instrumentalist'[Title/Abstract] OR 'singer'[Title/Abstract] OR 'vocalist'[Title/Abstract] OR 'theatre'[Title/Abstract] OR 'actor'[Title/Abstract] OR 'actress'[Title/Abstract] OR 'circus performer'[Title/Abstract] OR 'circus arts'[Title/Abstract] OR 'performing arts'[Title/Abstract] OR 'occupational performer'[Title/Abstract])) AND ('elite' OR 'professional' OR 'full-time student' OR 'pre-professional' OR 'non-recreational performer*')) AND ('screening' OR 'predictor' OR 'predictive tools' OR 'questionnaire*' OR 'survey*' OR 'outcome measures' OR 'risk assessment tools' OR 'assessment tools' OR 'assessment' OR 'inventory' OR 'evaluation tools' OR 'diagnostic tools' OR 'measur* tools')

Other filters: English language, adults, humans.

**Elsevier (Science Direct** - Limited search and no string combination option)

("psychosocial factors" OR "mental health" OR "depression" OR "burnout")AND("pain" OR "injury" OR "musculoskeletal pain")AND("performing arts" )

**SciVal (Scopus):**

Article title, abstract, keywords: 'psychosocial AND factors' OR 'mental AND health' OR 'anxiety' OR 'depression' OR 'catastrophizing' OR 'catastrophising' OR 'psychological OR stress' OR 'burnout' OR 'cognitive AND factors' OR 'stress' OR 'mental' OR 'emotional'

AND

Article title, abstract, keywords: 'pain' OR 'injury' OR 'musculoskeletal AND pain' OR 'chronic AND pain' OR 'musculoskeletal AND disorders' OR 'injury AND prevention' OR 'injury AND risk' OR 'wound' OR 'workplace AND injury'

AND

All fields: dancer OR dancing OR musician OR singer OR instrumentalist OR circus OR theater OR theatre OR actor OR actress OR performer

AND

All fields: elite OR pre-professional OR professional OR "NON-RECREATIONAL" OR "FULL-TIME STUDENT"

AND

All fields: 'screening' OR 'predictor' OR 'predictive AND tools' OR 'questionnaire*' OR 'survey*' OR 'outcome AND measures' OR 'risk AND assessment AND tools' OR 'assessment AND tools' OR 'assessment' OR 'inventory' OR 'evaluation AND tools' OR 'diagnostic AND tools' OR 'measur* AND tools'

**ProQuest (Performing Arts Periodical Database, Dissertations)**

summary('psychosocial factors' OR 'mental health' OR 'anxiety' OR 'depression' OR 'catastrophizing' OR 'catastrophising' OR 'psychological stress' OR 'burnout' OR 'cognitive factors' OR 'stress' OR 'mental' OR 'emotional') AND summary(('pain' OR 'injury' OR 'musculoskeletal pain' OR 'chronic pain' OR 'musculoskeletal disorders' OR 'injury prevention' OR 'injury risk' OR 'wound' OR 'workplace injury')) AND summary(('perform* artist' OR 'dancer' OR 'dancing' OR 'musician*' OR 'instrumentalist*' OR 'singer' OR 'vocalist' OR 'theatre' OR 'actor' OR 'actress' OR 'circus performer' OR 'circus arts' OR 'performing arts' OR 'occupational performer')) AND ('elite' OR 'professional' OR 'full-time student' OR 'pre-professional' OR 'non-recreational performer) AND ('screening' OR 'predictor' OR 'predictive tools' OR 'questionnaire*' OR 'survey*' OR 'outcome measure' OR 'risk assessment ' OR 'assessment tools' OR 'assessment' OR 'inventory' OR 'evaluation tools' OR 'diagnostic tools' OR 'measur* tools')

**SAGE:**

'psychosocial factors' OR 'mental health' OR 'anxiety' OR 'depression' OR 'catastrophizing' OR 'catastrophising' OR 'psychological stress' OR 'burnout' OR 'cognitive factors' OR 'stress' OR 'mental' OR 'emotional' AND 'pain' OR 'injury' OR 'musculoskeletal pain' OR 'chronic pain' OR 'musculoskeletal disorders' OR 'injury prevention' OR 'injury risk' OR 'wound' OR 'workplace injury' AND danc* OR 'perform* artist' OR 'dancer' OR 'dancing' OR 'musician*' OR 'instrumentalist*' OR 'singer' OR 'vocalist' OR 'theatre' OR 'actor' OR 'actress' OR 'circus performer' OR 'circus arts' OR 'performing arts' OR 'occupational performer' AND 'elite' OR 'professional' OR 'full-time student' OR 'pre-professional' OR 'non-recreational performer AND 'screen*' OR 'predict*' OR 'predictive tools' OR 'questionnaire*' OR 'survey*' OR 'outcome measure' OR 'risk assessment ' OR 'assessment tools' OR 'assessment' OR 'inventory' OR 'evaluation' OR 'diagnostic' OR 'measur* tools'

Since 1980 (no way to increase records on page)

**JSTOR**:

(((((ab:""perform" OR "singer" OR "danc*" OR "musician" OR "circus" OR "theatre"") AND ("stress" OR "anxiety" OR "catastroph*" OR "psychological")) AND ("pain" OR "injury" )) AND ("ELITE" OR "PROFESSIONAL" )) AND (ADULT))

**PEDRO:** As per webpage advanced search option: https://search.pedro.org.au/advanced-search
